# Understanding emotional and health indicators underlying the burnout risk of healthcare workers

**DOI:** 10.1101/2024.04.11.24305661

**Authors:** Elçin Güveyi, Garry Elvin, Angela Kennedy, Zeyneb Kurt, Petia Sice, Paras Patel, Antoinette Dubruel, Drummond Heckels

## Abstract

**Background:** Burnout of healthcare workers is of increasing concern as workload pressures mount. Burnout is usually conceptualised as resulting from external pressures rather than internal resilience and although is not a diagnosable condition, it is related to help seeking for its psychological sequelae.

**Objective:** To understand how staff support services can intervene with staff heading for burnout, it is important to understand what other intrapsychic factors that are related to it.

**Methods:** A diary tool was used by staff in a region of England to self monitor their wellbeing over time. The tool explores many areas of mental health and wellbeing and enabled regression analysis to predict which of the various factors predicted scores on the burnout item.

**Findings:** Burnout can be best explained with independent variables including *depression, receptiveness, mental wellbeing*, and *connectedness* (p<0.05) using a multiple linear regression model. It was also shown that 71% of the variance present in the response variable, i.e. burnout, explained by independent variables. There is no evidence found for multicollinearity in our regression models confirmed by both the Spearman Rank Correlation and the Variance Inflation Factor methods.

**Conclusion:** We showed how burnout can be explained using a handful number of factors including emotional and mental health indicators.

**Clinical implications:** The findings suggest a simple set of items can predict burnout and could be used for screening. The data suggests attention to four factors around social safeness, grounding and care in the self, hope and meaning and having sufficient energy could form the basis of attention in weelbeing programs.

## 1. Introduction

Burnout is defined as a psychological syndrome that emerges from work-related chronic stress (World Health Organisation, 2018). The experience was first described as three correlated yet distinct dimensions including emotional exhaustion, depersonalisation, and a reduced sense of self-actualisation (Freudenberger, 1975). The symptoms of burnout include feelings of energy depletion, negativism related to one’s job, detachment, low mood, a sense of ineffectiveness and lack of accomplishment (World Health Organisation, 2018). Burnout is more likely to happen when a job has a heavy workload, is understaffed or has conflicting and unrewarding work (Hert, 2020). An accumulation of research has identified a high prevalence of burnout among healthcare workers (West et al., 2016; Membrive-Jiménez et al., 2022; Pappa et al., 2021). Indeed, the health sector may be at greater risk of burnout than other professional fields as symptoms were observed in 37.9% of physicians as opposed to 27.8% in a population control sample (Shanafelt et al., 2012). The etiopathogenesis of burnout is multifactorial hence there are several sequential hypotheses that aim to understand the development process.

Freudenberger’s model of burnout was most recently revised into five consecutive stages that describe the developmental process (Freudenberger, 1982). The honeymoon phase specifies enthusiasm at the beginning of a job and is later followed by stagnation due to an absence of positive coping mechanisms when stressors of the job are introduced (Freudenberger, 1982). Work-life balance becomes distorted thus initiating the third stage of chronic stress characterised by feelings of failure, powerlessness, and incompetency (Freudenberger, 1982). Apathy develops with feelings of hopelessness and disparity leading to habitual burnout where one may seek help and intervention (Freudenberger, 1982). This description of burnout employs a causal sequence when coping with stress which may be preventable by greater control. The Job Demand-control model proposes that the risk of burnout may be implemented by an imbalance between the height of strain and the level of control in a work environment (Karasek, 1979). Low-high strain jobs can be represented through work rate, availability, time pressure, and difficulty of tasks (Karasek, 1979). The level of control in an employee refers to their freedom to organise and manage the workload (Karasek, 1979). Hence, there are several highly demanding jobs that impose stress, but burnout may only appear depending on the level of control and personal attitude of the employee.

COVID-19 magnified burnout in front-line workers as they were expected to work in stressful work environments with chronic underinvestment in the public health infrastructure, escalating workload burdens, inadequate support, and moral injury from being unable to provide essential care to patients. Emotional exhaustion and depersonalisation were higher in 2021 than observed in 2020,2017,2014 and 2011 (Shanafelt et al., 2022). Work-life integration declined significantly by 16.1% from 2020 to 2021 amongst physicians. Furthermore, the lack of personal protective equipment, routine testing, staff shortages, inadequate training, rapidly changing guidelines, and risk of transmission of infections to friends and family were contributory factors of burnout for healthcare workers (Vindrola-Padros et al., 2020; Nyashanu et al., 2020; Papoutsi et al., 2020). This significant decline in healthcare workers’ well-being since the pandemic suggests that worksite features are fundamental in promoting burnout.

Adjacent to worksite features, the decision latitude of healthcare workers may have increased the likelihood of burnout as Eder and Meyer (2022) propose the concept of self-endangering work behaviour defined as actions that aim to deal with work-related demands yet simultaneously elicit health problems. This qualitative study found that self-endangering behaviour was an essential precursor of nurses’ burnout and was based on the altruistic attitude of boosting one’s self-esteem by helping others. These behaviours may include extending work time, reducing recovery time and working overtime. The process of burnout may therefore develop through this vicious cycle and impact one’s physical and mental health. A systematic review of twelve studies found that the presence of sleep disorders was profound in nurses with a higher level of burnout (Membrive-Jiménez et al., 2022). Additionally, there are significant associations between burnout and depression (Koutsimani et al., 2019). Advanced ages and longer years of work intensified the relationship between depression and burnout which may suggest that repeated and negative experiences are necessary for burnout to develop into overlapping symptoms of depression (Meier et al., 2022).

The aim of this study was to explore the correlations between various wellbeing and mental health factors and burnout, which would provide a clearer idea of the meaning and struggles of staff reporting this experience. By modelling the concept of burnout in this local population it was hoped that clearer routes to healing would emerge.

## 2. METHODS

### 2.1. Diary and User Interface

‘My Personal Wellbeing’ is a web based self-monitoring and wellbeing screening diary facility for the Covid-19 pandemic and beyond. Participants were encouraged to reflect and provide descriptions of experience and their sense of wellbeing using a self-reflection diary based on a holistic integrative wellbeing model. The diary aims to help users identify and understand patterns in a number of aspects of their wellbeing to support them in managing and maintaining good wellbeing. This proactive insight allowed for forward planning of the NHS’ regional Staff Wellbeing Hub Services for The North East and North Cumbria region to ensure suitable and timely interventions were available to staff at the time they needed the most. Data is collected using both quantitative and qualitative methods to enable objective statistical and sentiment analysis of the data.

The online diary consists of 26 wellbeing factors that are rated on a slider scale from positive to negative. Individual responses are confidential and therefore users can feel confident about being honest with their responses. Its purpose allows self monitoring of wellbeing. It also gives a service that supports staff some aggregated public health information that can be used to meet their needs. Detailed information about the process of diary design was published in the previous study (Elvin et al., 2023).

Table 1 describes the question codes and phrases shown to the participants on the online diary but only for those that were significant in the prediction model. Other items included sleep quality, usage of drugs and alcohols, moral injury, value to life, emotional and physical wellbeing were excluded after the regression analysis due to not having significant p-values.

**Table 1.**
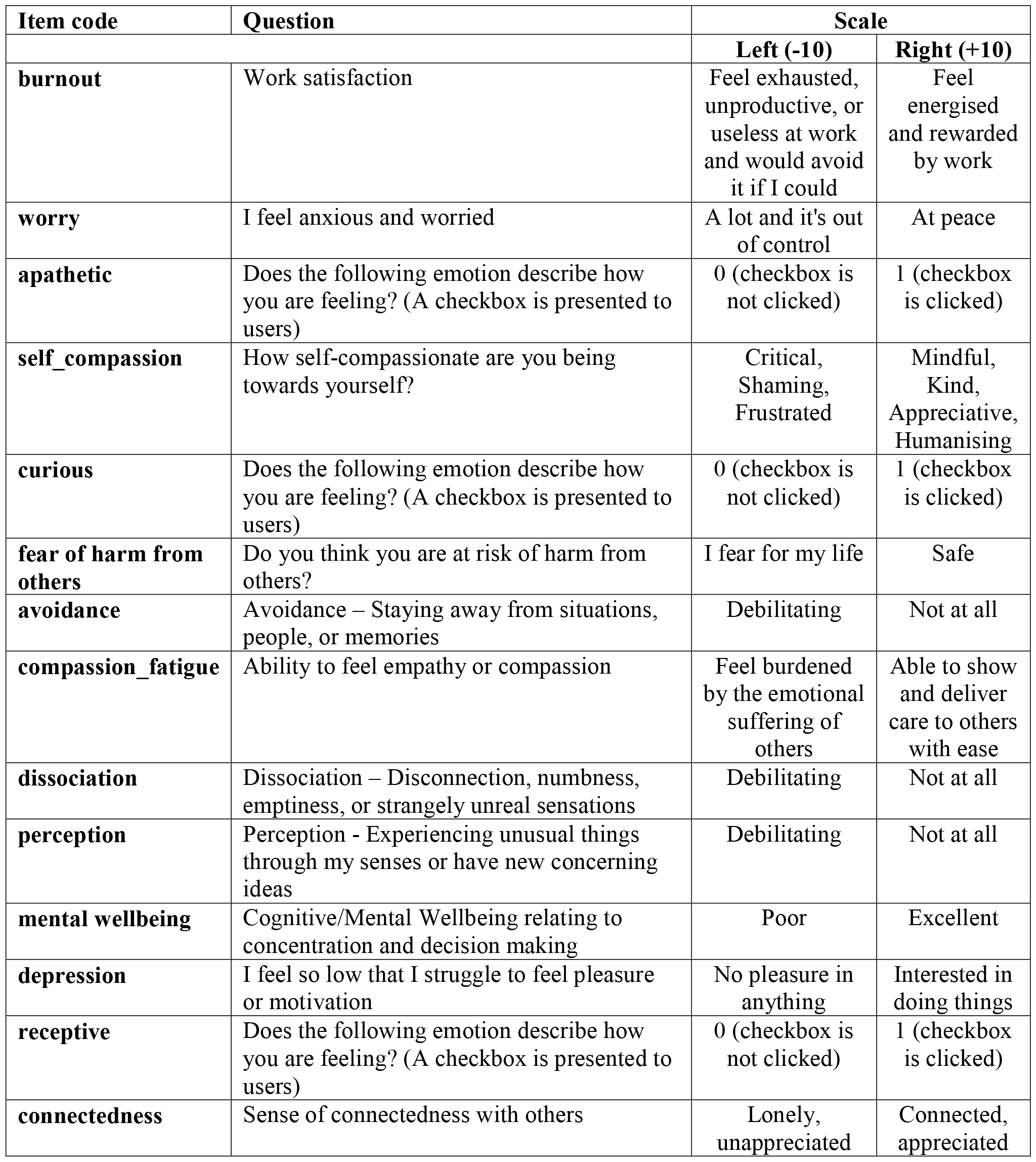
Questionnaire item codes and the related questions.

| Item code | Question | Scale |  |
| --- | --- | --- | --- |
|  |  | Left (-10) | Right (+10) |
| **burnout** | Work satisfaction | Feel exhausted, unproductive, or useless at work and would avoid it if I could | Feel energised and rewarded by work |
| **worry** | I feel anxious and worried | A lot and it's out of control | At peace |
| **apathetic** | Does the following emotion describe how you are feeling? (A checkbox is presented to users) | 0 (checkbox is not clicked) | 1 (checkbox is clicked) |
| **self_compassion** | How self-compassionate are you being towards yourself? | Critical, Shaming, Frustrated | Mindful, Kind, Appreciative, Humanising |
| **curious** | Does the following emotion describe how you are feeling? (A checkbox is presented to users) | 0 (checkbox is not clicked) | 1 (checkbox is clicked) |
| **fear of harm from others** | Do you think you are at risk of harm from others? | I fear for my life | Safe |
| **avoidance** | Avoidance – Staying away from situations, people, or memories | Debilitating | Not at all |
| **compassion_fatigue** | Ability to feel empathy or compassion | Feel burdened by the emotional suffering of others | Able to show and deliver care to others with ease |
| **dissociation** | Dissociation – Disconnection, numbness, emptiness, or strangely unreal sensations | Debilitating | Not at all |
| **perception** | Perception - Experiencing unusual things through my senses or have new concerning ideas | Debilitating | Not at all |
| **mental wellbeing** | Cognitive/Mental Wellbeing relating to concentration and decision making | Poor | Excellent |
| **depression** | I feel so low that I struggle to feel pleasure or motivation | No pleasure in anything | Interested in doing things |
| **receptive** | Does the following emotion describe how you are feeling? (A checkbox is presented to users) | 0 (checkbox is not clicked) | 1 (checkbox is clicked) |
| **connectedness** | Sense of connectedness with others | Lonely, unappreciated | Connected, appreciated |

### 2.2. Participants

The participants for this study were NHS and social care professionals, including nurses, doctors, and administrators. They were invited to complete an online diary with 26 questions about personal wellbeing. The recruitment period for this study started on the 4th January 2021 and completed on the 20th April 2022. To enter the study and gain access to the diary, participants were first required to register and confirm their consent for their data to be processed by clicking a checkbox. Participation was on a voluntary basis. Since the aim of the study was to investigate change in wellbeing over time, individuals who completed the diary only once were excluded from the analysis, leaving a total of 73 participants and 219 entries. The distribution of entries over the period of this study is shown in Supporting Table 1.

### 2.3. Regression Analysis

Regression analysis is a predictive model trained to understand whether there is an association between a target variable and a set of independent variables. We can also investigate which explanatory variables are the most relevant ones to the dependent variable while predicting the relationship between them. Since we have dependent and independent variables the find the correlation with burnout, regression analysis is the best way to reveal the linear relationship between the target variable and the other factors. In the present study, multiple linear regression analysis was performed to explore the related factors to burnout. Two-stage regression analysis was planned: the first stage aims to find factors that are related to burnout (p-value under 0.05), and the second stage aims to find high-significant items over the regression model established with these related factors. All statistical analyzes was conducted using R version 4.2.1 in RStudio version 2022.07.1+554. R package *stats* was used with *lm* function to carry out regression models.

Assessment of the regression model performance was made by employing the R-Squared (R2) metric. This metric shows the proportion of the variance that explained by independent variables in a regression model (Helland, 1987; Granger et. al, 1976). Validation of the R-Squared values was fulfilled with the 5-Fold Cross Validation method with ten times repeats. The dataset was split into five evenly distributed parts and in every iteration one of these parts separated for testing the model and the other parts were used for training and constructing the regression model. In each iteration, the test dataset was fitted to the constructed model and the predicted values were obtained with *predict* function in the *stats* package. R-Squared metric was calculated for each iteration with *R2* function from caret package in R. After five iterations, average R2 values was obtained. We repeated cross validation process for 10 times to ensuring model’s performance consistency.

### 2.4. Analysis of Covariances

ANCOVA (Analysis of Covariance) is composed of regression and ANOVA (Analysis of Variances) analyses. ANCOVA analysis helps to find out how a variable, called covariate, affects model prediction success on a continuous dependent variable with grouping based on a categorical independent variable (Fisher, 1970). While performing ANCOVA, one of the independent variables should be categorical, whilst the dependent variable and covariate should be continuous variables. After obtaining regression results, variables that are likely to be covariants will be checked with ANCOVA. *ancova* function from R package *jmv* will be used to perform analysis of covariances.

### 2.5. Check for Multicollinearity

In multiple linear regression models, one of the important factors threatening model reliability is multicollinearity existence. Multicollinearity is defined as strong correspondence between two or more independent variables in a regression model (Belsley et al., 1980). This strong relationship can causes the regression model to perform misleading results. Correlation analysis and calculation of Variance Inflation Factor (VIF) are well-known ways to determine the presence of multicollinearity. Correlation analysis on the regression model’s variables was performed with Spearman Rank Correlation. In RStudio, the *stats* package’s *cor* function was used and set the “*method=spearman*” to obtain the correlation scores and the *gplots* package’s *heatmap.2* function was used to visualize the relationship between variables based on these scores. Variance Inflation Factor (VIF)(Marquaridt, 1970) is another important metric for detecting dependency between variables. It is widely used in the literature to validate regression models (Gregorich et al., 2021; Vu et al., 2015). VIF score calculation was performed with *car* package’s *vif* function in R.

## 3. RESULTS

### 3.1. Findings from the Regression Model

The dataset used in the study consists of 37 questionnaire items for wellbeing and symptom-related questions. The burnout item asked diary users to rate their extent of work satisfaction by positioning a slider between two poles of ‘Feel energised and rewarded by work’ to ‘Feel exhausted, unproductive, or useless at work and would avoid it if I could’. In the multiple linear regression model, “burnout” was selected as target variable and the rest of the other 36 items included as independent variables of the model. At first step, variables that have a p-value below the significance level of 0.05 were selected as informative variables and then the regression model was reconstructed with these items. Table 2 shows the burnout-targeted regression model statistical summary results. Our multiple linear regression analysis highlighted which factors from many are the important ones in prediciting the target variable’s vaule. A combination of wellbeing factors could predict 70% of the variation in burnout scores.

**Table 2.** Multiple linear regression model summary statistics in predicting burnout.

|  | *B* | *SE* | *t-value* | *p-value* |
| --- | --- | --- | --- | --- |
| **worry** | 0.1208282 | 0.06547753 | 1.8453386 | 0.0664306 |
| **apathetic** | -2.4685710 | 0.63340665 | -3.8972925 | 0.0001317* |
| **self compassion** | 0.2247986 | 0.06579436 | 3.4166841 | 0.0007643* |
| **curious** | 4.7569654 | 2.15384560 | 2.2085916 | 0.0283110* |
| **fear of harm from others** | -0.1480708 | 0.05596174 | -2.6459295 | 0.0087786* |
| **avoidance** | 0.1385615 | 0.06643903 | 2.0855442 | 0.0382577* |
| **compassion fatigue** | 0.1943516 | 0.05414616 | 3.5893893 | 0.0004147* |
| **dissociation** | -0.1090891 | 0.05820600 | -1.8741907 | 0.0623267 |
| **perception** | 0.1004212 | 0.05532519 | 1.8151084 | 0.0709686 |
| **mental wellbeing** | 0.2975105 | 0.07005322 | 4.2469213 | 0.0000329* |
| **depression** | 0.3202704 | 0.06071517 | 5.2749640 | 0.0000003* |
| **receptive** | -7.3354672 | 1.68817551 | -4.3452042 | 0.0000219* |
| **connectedness** | -0.2640955 | 0.06288644 | -4.1995619 | 0.0000399* |

Note: *B* stands for Beta values and *SE* represents the standart error.
\* Statistically significant variables with a p-value below the 0.05

Significance level 0.05 was selected as threshold and the items have p-values below this value as marked as significant items of the regression model in Table 2. Items entitled *apathetic, self compassion, compassion fatigue, mental wellbeing, depression, receptive*, and *connectedness* seem highly related (p-value under the 0.001) to *burnout*.

In order to validate the regression model outcomes, R-Squared metric was calculated on the independent test dataset with 5-Fold Cross Validation. After ten times repeated cross validation on the dataset, average R2 values were obtained as follows (Table 3).

**Table 3.** Average R-Squared values for each iteration.

|  | **Iterations** |  |  |  |  |  |  |  |  |  |
| --- | --- | --- | --- | --- | --- | --- | --- | --- | --- | --- |
|  | **1** | **2** | **3** | **4** | **5** | **6** | **7** | **8** | **9** | **10** |
| **Average 5-Fold** | 0.703 | 0.709 | 0.722 | 0.712 | 0.710 | 0.703 | 0.720 | 0.718 | 0.695 | 0.711 |
| **Average** | **0.7103 ± 0.01** |  |  |  |  |  |  |  |  |  |

The mean value of R2 was found to be 0.71 after being repeated 10 times. This shows that more than 70% of the variance in burnout as described by extent of work satisfaction can be explained by the independent variables.

### 3.2. ANCOVA (Analysis of Covariance)

Self compassion and compassion fatigue were found to be highly significant predictors in our regression model and they are known to be conceptually related (Hashem & Zeinoun, 2020, Beaumont et al, 2015). We checked whether any covariates exist in our linear model using ANCOVA, and focused on this highly relevant and significant predictors of the burnout model. In ANCOVA one of these variables is expected to be a categorical variable and the rest of the predictors to remain as continuous variables, so is the dependent variable. A univariate analysis (i.e. histogram) for *self compassion* and *compassion fatigue* was performed to determine which item is more suitable for categorization. *self-compassion* is normally distributed around a zero-mean (see Supp Fig S1). But some participants might not use the sliding bar and keep the diary score at the default value, i.e. zero, which corresponds to the mod of the histogram of this item. When the peak bar at the point zero is omitted, self-compassion reflects an approximate uniform distribution, which evidences that it is more suitable for discretization because of the almost equal distribution of the samples across different categorical values of this variable.

Hence, *self compassion* was reshaped as a categorical variable: values between −10 and −4 are named as “low self compassion” (high self criticism), −3 and 3 range was named as “medium”, 4 and 10 range was named as “high self compassion”. ANCOVA model’s dependent variable was assigned as the diary item of *burnout* in *ancova* function from *jmv* package in R. Table 4 shows the ANCOVA model’s statistical summary results.

**Table 4.**
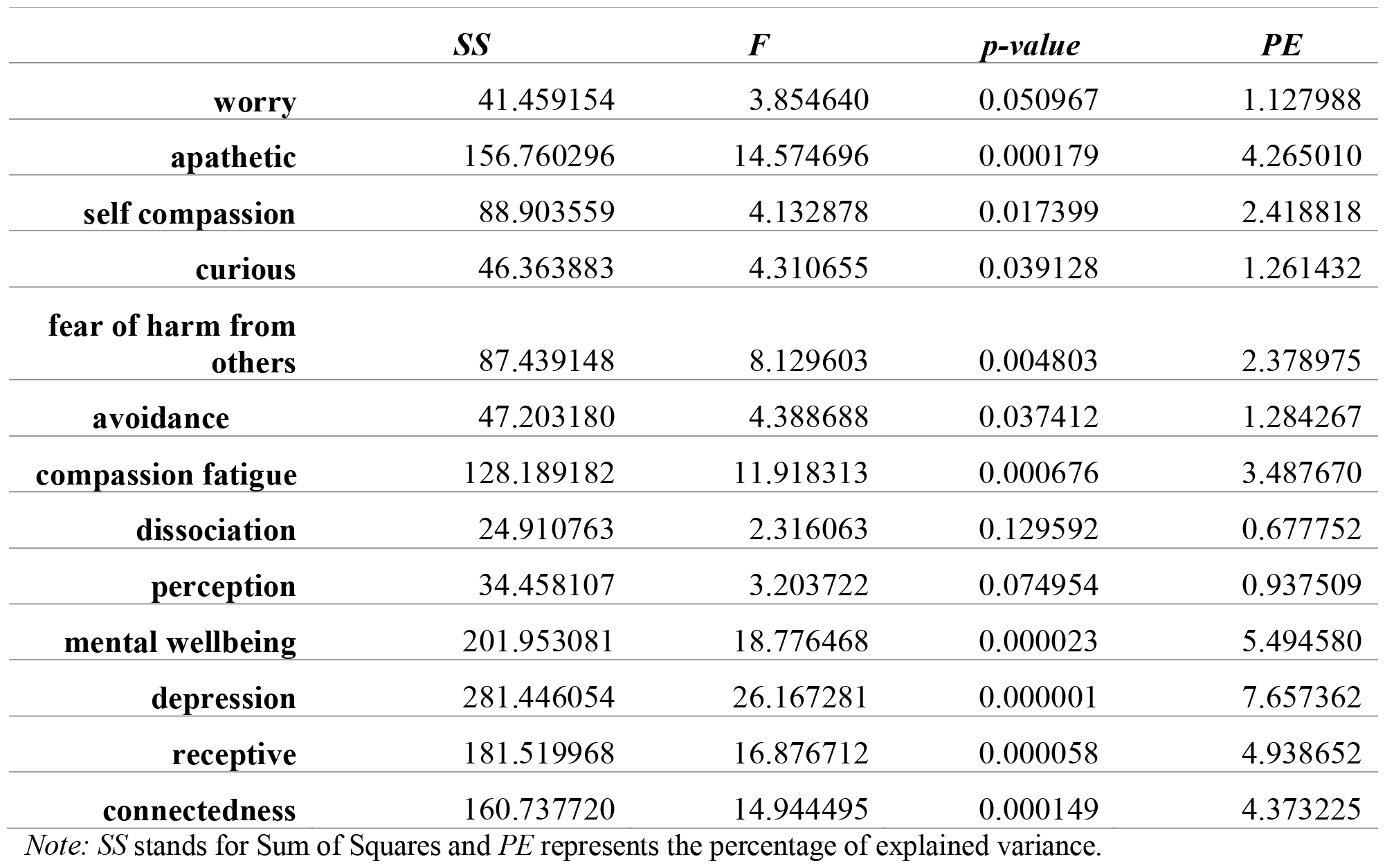
Statistical summary results for ANCOVA analysis.

|  | *SS* | *F* | *p-value* | *PE* |
| --- | --- | --- | --- | --- |
| **worry** | 41.459154 | 3.854640 | 0.050967 | 1.127988 |
| **apathetic** | 156.760296 | 14.574696 | 0.000179 | 4.265010 |
| **self compassion** | 88.903559 | 4.132878 | 0.017399 | 2.418818 |
| **curious** | 46.363883 | 4.310655 | 0.039128 | 1.261432 |
| **fear of harm from others** | 87.439148 | 8.129603 | 0.004803 | 2.378975 |
| **avoidance** | 47.203180 | 4.388688 | 0.037412 | 1.284267 |
| **compassion fatigue** | 128.189182 | 11.918313 | 0.000676 | 3.487670 |
| **dissociation** | 24.910763 | 2.316063 | 0.129592 | 0.677752 |
| **perception** | 34.458107 | 3.203722 | 0.074954 | 0.937509 |
| **mental wellbeing** | 201.953081 | 18.776468 | 0.000023 | 5.494580 |
| **depression** | 281.446054 | 26.167281 | 0.000001 | 7.657362 |
| **receptive** | 181.519968 | 16.876712 | 0.000058 | 4.938652 |
| **connectedness** | 160.737720 | 14.944495 | 0.000149 | 4.373225 |

Note: *SS* stands for Sum of Squares and *PE* represents the percentage of explained variance.

The results from Table 4 show that *apathetic, compassion fatigue, mental wellbeing, depression, receptive*, and *connectedness* variables are highly significant and hold the important amount of variance. All the variables except *worry* and *perception* have the p-value below 0.05. This means that these variables affect the prediction outcome and it’s the right choice to include these ones in the regression model. We concluded that the *self compassion* and *compassion fatigue* items have statistically significant p values indicating that compassion fatigue is a covariate for the predictor categorical variable self-compassion within our linear model.

### 3.3. Correlation Analysis and VIF Scores

To see whether there is multicollinearity between the variables in the regression model, correlation analysis and VIF score calculation were performed. The heatmap created with correlation scores and the clustered items is shown in Figure 1.

**Figure 1.**
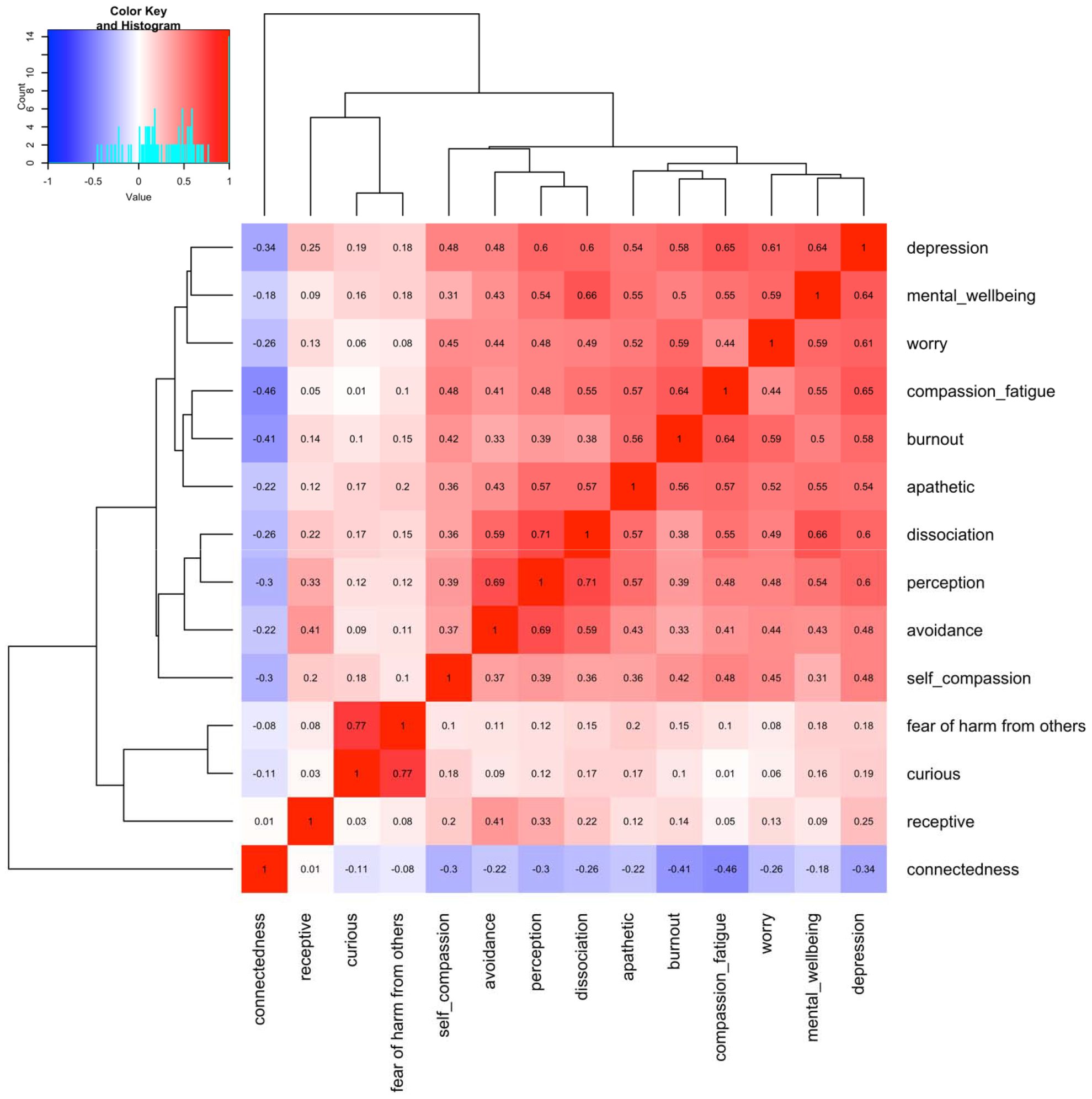
Correlation scores for variables in the regression model.

The results that can be inferred from the heatmap are as follows:

- *curious* and *receptive* have the highest correlation in the heatmap with coefficient of 0.77
- *dissociation* and *avoidance* have correlation higher than 0.7 (r=0.71).
- *dissociation* and *perception* have almost 70% correlation (r=0.69).
- *avoidance* and *worry* have a correlation coefficient of 0.66
- *burnout* and *mental wellbeing* have a correlation coefficient of 0.64
- there seem to be three distinct clusters occured, (i) a group consists of burnout, self compassion, mental wellbeing, connectedness, worry, depression; (ii) avoidance, dissociation, perception and compassion fatigue seem to be relevant to one another; meanwhile (iii) fear of harm from others, receptiveness, and curiousity are groupped together. Apathetic seems to remain further away from the rest of the items, and it has a weak to moderate negative correlation with the rest.

Correlation scores higher than 0.8 show that there can be multicollinearity between the variables and one of them can be kept and the rest can be removed from the model. We concluded that none of the independent variables had a correlation higher than 0.77. Thus, we hypothesised that there is no need to exclude any of the variables from the model based on the correlation scores.

Another important multicollinearity checking method Variance Inflation Factor (VIF) was applied to the regression model. VIF score can have values starting from 1. Score 1 shows that there is no significant correlation between the variables of the linear model. Interval between 1 and 5 indicates that the variables are moderately correlated. The VIF score greater than 5 means the correlation is high (Chatterjee et al., 2006). VIF metric was calculated for each independent variable in the burnout-targetted regression model (Table 5).

**Table 5.** VIF scores of the independent variables in burnout targetted regression model.

| Item | VIF score |
| --- | --- |
| worry | 2.570105 |
| apathetic | 1.338757 |
| self compassion | 2.107946 |
| curious | 2.569246 |
| fear of harm from others | 1.325771 |
| avoidance | 3.006439 |
| compassion fatigue | 1.496584 |
| dissociation | 2.888239 |
| perception | 2.130852 |
| mental wellbeing | 2.284351 |
| depression | 2.709448 |
| receptive | 2.581232 |
| connectedness | 2.173744 |

According the results shown in Table 5, all VIF scores for the independent variables are between 1 and 3. Which means there is no significant relationship between the variables and the regression model is valid. Thus, we don’t need to exclude any variable from the model.

## 4. DISCUSSION

Burnout, as assessed by a simple question about the extent of exhaustion or energy for work, could be significantly predicted by a small subset of wellbeing items. These were how compassionate the person is being to themselves, ability to feel empathy or compassion for others, ability to concentrate and make decisions, low mood that limits pleasure, sense of connectedness to others, *apathetic, receptive, curious*. The model provides textural information on the concept of burnout that can be nebulous. It has been defined as a state of exhaustion characterised by a range of emotions, lack of concentration and decision making, insomnia and other physical health issues, a sense of alienation or isolation and behaviours such as using alcohol or withdrawal or lack of empathy. This practice based model specifies that within this particular health and care population, some of those factors such as sleep, alcohol, irritability, physical wellbeing, were not significant. However the main factors found here aligned with the previous literature. Such wellbeing factors can have a significant impact on capacity to function and on retention of good staff (The Society of Occupational Medicine).

This cluster of burnout items could form the basis of screening at work by using the small number of simple questions in Table 1. Early engaging mindful awareness of burnout as it creeps it could then lead to early or preventative interventions. Some interventions may be aimed at the staff themselves. Mindful Self Compassion training has been found to be helpful (Kotera and Gordon, 2021). Opportinities to connect with others in ways that bring joy and energy may help (Matthew et al.,2022). However burnout is a consequence of working conditions. Modelling compassionate leadership and lived experience is key (Panagioti et al 2016). The Heath and Safety Executive has management standards that covers six areas that mitigate against employee burnout such as workload, choice, meaningful work, fairness, supportive teamwork, and sufficient positive recognition for staff efforts (Great Britain Health and Safety Executive, 2019). Peer support (Johnson, 2023) and Schwartz rounds (Hogan et al.,2022) have been interventions that fall between the macro and the micro interventions. Both show they contribute towards a healthy work culture.

On the basis on the four clusters identified, diagram in Supp Fig S2 suggests that wellbeing can be characterized by four overarching areas. One is that of social safeness. The felt sense of belonging and safety within a team or community. Another is the aliveness within the self: Feel positive things for oneself and not be troubled by unusual perceptual experiences. A third area was related to cognitive aspects of hope, meaning and capacity to be focused and directed. The final cluster related to empathy for others, energy for work and capacity to be engaged. These domains could inform the structure of wellbeing awareness or leadership programs and suggest that attention to social factors, cognitions as reactions to working conditions compassion would form the basis of cultural wellbeing.

## Data Availability

All data produced in the present study are available upon reasonable request to the authors

## Ethics declarations

This study has been approved by the Ethics Committee of the Faculty of Engineering and Environment at Northumbria University, Newcastle upon Tyne, with an approval reference number 23709. Informed consent was obtained from all participants or their legal guardian(s). Participants were provided details about the study and what data/information will be collected from them and were asked to agree to take part before any data was collected. All methods were conducted in accordance with relevant guidelines and regulations.

